# Modeling the COVID-19 dissemination in the South Region of Brazil and testing gradual mitigation strategies

**DOI:** 10.1101/2020.07.02.20145136

**Authors:** Rafael M. da Silva

## Abstract

This study aims to understand the features of the COVID-19 spread in the South Region of Brazil by estimating the Effective Reproduction Number (ERN) *ℛ*_*e*_ for the states of Paraná (PR), Rio Grande do Sul (RS), and Santa Catarina (SC). We used the SIRD (Susceptibles-Infectious-Recovered-Dead) model to describe the past data and to simulate strategies for the gradual mitigation of the epidemic curve by applying non-pharmacological measures. Besides the SIRD model does not include some aspects of COVID-19, as the symptomatic and asymptomatic subgroups of individuals and the incubation period, for example, in this work we intend to use a classical and easy to handle model to introduce a thorough method of adjustment that allows us to achieve reliable fitting for the real data and to obtain insights about the current trends for the pandemic in each locality. Our results demonstrate that for localities for which the ERN is about 2, only rigid measures are efficient to avoid overwhelming the health care system. These findings corroborate the relevance of keeping the value of *ℛ*_*e*_ below 1 and applying containment measures early.

## 1. Introduction

On March 11^th^, 2020, the infectious disease caused by the most recently discovered Coronavirus (SARS- CoV2), the Coronavirus Disease COVID-19, was characterized as a pandemic by the World Health Organization (WHO) [1]. The new Coronavirus astonished the world with its high infectivity, which allowed the quick spread of the COVID-19 across all continents. Over the last months, researchers have concentrated their efforts to obtain answers and to propose efficient measures to avoid an even greater catastrophe. Despite no vaccine has been developed yet, to understand the process of dissemination of the virus [2] and to propose universal strategies to flatten the epidemic curve through social distance protocols [3, 4], for example, were fundamental achievements.

After starting in China and affecting several countries in Europe, as well as the United States of America (USA), currently, South America is the epicenter of the COVID-19 pandemic. Until June 20^th^, 2020, in Brazil, Peru, and Chile, the three most affected countries of such continent, more than 1.5 million people were infected. In Brazil, due to its continental dimension, the COVID-19 spread across its 27 states (considering the federal district) heterogeneously, as shown recently [5]. Some states in the North, Northeast, and Southeast Brazilian regions were the most affected. Considering only São Paulo and Rio de Janeiro, located in the Southeast Region, and Ceará and Pará, belonging to the Northeast and North regions, respectively, we get almost 46% of the cumulative number of confirmed cases and 63% of the deaths caused by COVID-19 in the whole country.

On the other hand, the South and Central-West regions, for a while, remain with a low incidence of cases. Together, these two regions concentrate about 10% of the cumulative number of confirmed cases and about 5% of the deaths in Brazil. Because such disparity when comparing different regions, large countries as Brazil, in which cultural, climatic, and per capita income differences take place, for example, are studied so that we can find out which aspects indeed influence the virus spreading. The main focus of the studies already carried out is on the most affected regions by the epidemic [5, 6, 7, 8]. In this work, however, we propose to analyze the current situation in the Brazilian South Region, which is composed of the states of Paraná (PR), Rio Grande do Sul (RS), and Santa Catarina (SC). The population, the cumulative number of confirmed cases, and the number of deaths caused by COVID-19 in these three states considering the data until June 20^th^, 2020, are presented in Table 1.

**Table 1.**
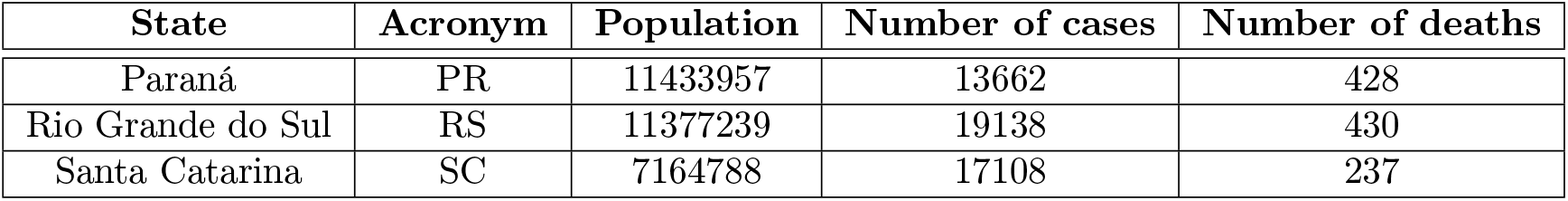
Table with the population, the cumulative number of confirmed cases, and the number of deaths caused by COVID-19 in the states of Paraná (PR), Rio Grande do Sul (RS), and Santa Catarina (SC) considering the data until June 20^th^, 2020. Data obtained from the Ref. [14].

| State | Acronym | Population | Number of cases | Number of deaths |
| --- | --- | --- | --- | --- |
| Paraná | PR | 11433957 | 13662 | 428 |
| Rio Grande do Sul | RS | 11377239 | 19138 | 430 |
| Santa Catarina | SC | 7164788 | 17108 | 237 |

To better understanding the dissemination of an infectious disease in a specific region, the most common approach is to use the different epidemiological models to obtain short-term or long-term forecasts and to simulate mitigation strategies [3, 4, 9, 10, 11, 12, 13]. In this study, we used the SIRD (Susceptibles-Infectious-Recovered-Dead) model, which is introduced in Section 2 together with the methodology used to adjust the model to the real data. In Section 3, we present our results, which are divide into three parts: (*i*) an estimate of the Effective Reproduction Number (ERN) *ℛ*_*e*_ for each state; (*ii*) projections for the cumulative number of confirmed cases and active infected people considering different scenarios; and (*iii*) mitigation strategies with the gradual reduction of *ℛ*_*e*_ to avoid the health care systems collapse. Our conclusions are presented in Section 4.

## 2. The SIRD model

Since the remarkable work published in 1927 by W. O. Kermack and A. G. McKendrick [15], the epidemiological models are taken to capture the essential features of the course of an epidemic. According to Daley and Gani [16], the use of epidemiological models has three main goals: (*i*) to understand better the mechanism by which diseases spread, (*ii*) to predict the future course of the epidemic, and (*iii*) to simulate strategies which allow us to control the spread of the disease. However, to achieve these three goals, first of all, our model must be able to describe the real data from the past [16].

Based on this premise, in this work, we used the SIRD model to describe the real data from the past and to project future scenarios for the COVID-19 pandemic. In the SIRD model, the total population *N* of each locality is divided into subgroups of susceptible (*S*(*t*)), infectious (*I*(*t*)), recovered (*R*(*t*)), and dead (*D*(*t*)) individuals, as shown in Fig. 1. These variables are functions of time *t* and the condition *N* = *S*(*t*) + *I*(*t*)+*R*(*t*)+*D*(*t*) is always satisfied. In our simulations, we considered the total population *N*, given in Table 1 for each state, a fixed number. The reason for this assumption is that we can safely ignore births and deaths to make predictions over a short period (a few months) [17].

**Figure 1.**
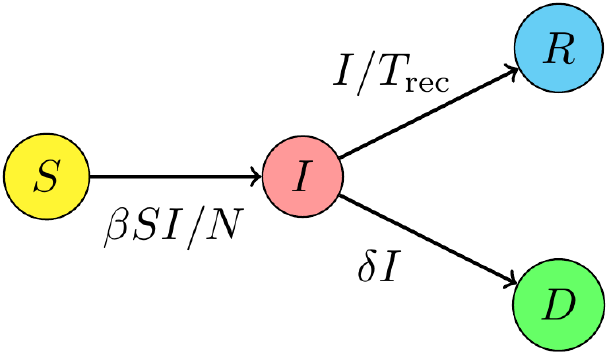
Schematic representation of the dynamics of the subgroups with susceptibles *S*, infectious *I*, recovered *R*, and dead *D* individuals.

The set of Ordinary Differential Equations (ODEs) that describes the dynamics of each subgroup is the following [5, 11, 15, 18]: 

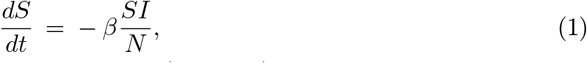

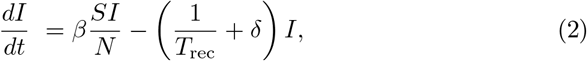

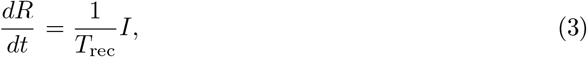

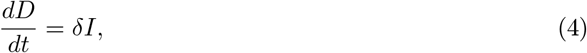

with initial conditions [*S*(*t*_0_), *I*(*t*_0_), *R*(*t*_0_), *D*(*t*_0_)]. In our simulations, we considered the initial number of susceptibles individuals as *S*(*t*_0_) = *N* −*I*(*t*_0_) −*R*(*t*_0_) − *D*(*t*_0_), with *R*(*t*_0_) = 0. We obtained *I*(*t*_0_) and *D*(*t*_0_) from the real data. The parameter *β* is the contact rate, the average number of adequate contacts (sufficient for transmission) of a person per unit time. Then, (*βI/N*) is the average number of contacts of one susceptible individual with infectious individuals per unit time, and (*βI/N*)*S* is the number of new cases per unit time due to *S* susceptibles [19, 20, 21]. Since the incidence of new infections depends on the prevalence (or “frequency”) *I/N* of the disease in the population, the term (*βI/N*)*S* is sometimes called *frequency-dependent incidence* (also called standard incidence or density-independent transmission) [22, 23]. 1*/T*_rec_ is the recovery rate, with *T*_rec_ being the recovery time (or duration of the infection), and the mortality (or death) rate is *δ* (number of people that die per unit of time comparing with the number of active infected individuals).

### 2.1. Adjusting the parameters to the real data

The SIRD model is very simple and does not consider the subgroups of symptomatic and asymptomatic individuals or the period in which a person is not infectious during incubation (until a few days before the onset of symptoms), as other more robust models [3, 9, 10, 13, 24]. However, the purpose of this work is to show how simple models, which are very important as conceptual models, can reproduce the real data accurately when an in-depth adjustment is performed to determine the best parametric set.

The parameters *β, T*_rec_, and *δ* can change daily and must be adjusted so that the model describes the real data from the past and allows us to make reliable projections for the future. To achieve the best adjustment, we divided the time series in Epidemiological Weeks (EPI weeks), which are, by international convention, count from Sunday to the next Saturday. The aim is to find the combination of *β*(*k*), *T*_rec_(*k*), and *δ*(*k*) that better reproduces the course of the pandemic in each EPI week number *k*, in a specific locality. Splitting the time series into intervals of seven days, with each day being an experimental data, we satisfied the required number of 2*r* + 1 experiments that must be considered to obtain the relevant information of the *r* parameters [25] (in this case, *r* = 3).

The adjustment procedure consisted of minimizing the Root Mean Square Error (RMSE) between the real data and the model predictions. We defined Δ_*C*_ as the RMSE calculated between the cumulative number of infected people *C*(*t*) = *I*(*t*) + *R*(*t*) + *D*(*t*) obtained by the model in each day and the real data, and Δ_*D*_ as the RMSE calculated between the number of deaths *D*(*t*) obtained by the model in each day and the real data. Thus, the first step of the numerical procedure was to perform simulations varying *β ∈* [0.0, 0.3] with a step of 10^−2^, *T*_rec_ *∈* [7, 21] with a step of 10^−1^, and *δ∈* [5 *×*10^−4^, 10^−2^] with a step of 5*×* 10^−5^ and to find the parametric set (*β*(*k*), *T*_rec_(*k*), *δ*(*k*)) that minimizes the quantity 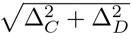 in each EPI week number *k*. Once Δ_*C*_≫ Δ_*D*_ (due to the magnitude of the data), the next step was to vary the parameter *δ*(*k*) again keeping fixed the parameters *β*(*k*) and *T*_rec_(*k*) found in the previous step, aiming to find a new parametric set (*β*(*k*), *T*_rec_(*k*), *δ*(*k*)) that minimizes Δ_*D*_. After this, we kept fixed the death rate and varied one more time *T*_rec_(*k*) and *β*(*k*), one after the other, to minimize the RMSE Δ_*C*_. Depending on the behavior of the real data, in the last step, *T*_rec_(*k*) and *β*(*k*) can remain unchanged.

We picked up the range 7-21 days for the recovery time because it covers a reliable estimate for the period in which the people might remain infectious [26, 27, 28, 29]. On the other hand, for the states of PR, RS, and SC, a variation from 0.05% to 1% for the death rate *δ* is a satisfactory range [14, 30]. To obtain better results, the first day considered on the fitting procedure is the next Sunday after the first death in each locality. For this reason, the adjustment for the three states begins in the EPI week number 14, as shown in Fig. 2. In this figure, the real data for *C*(*t*) and *D*(*t*) are displayed together with the model predictions, showing that the methodology proposed in this work can describe the natural oscillations of real time-series.

**Figure 2.**
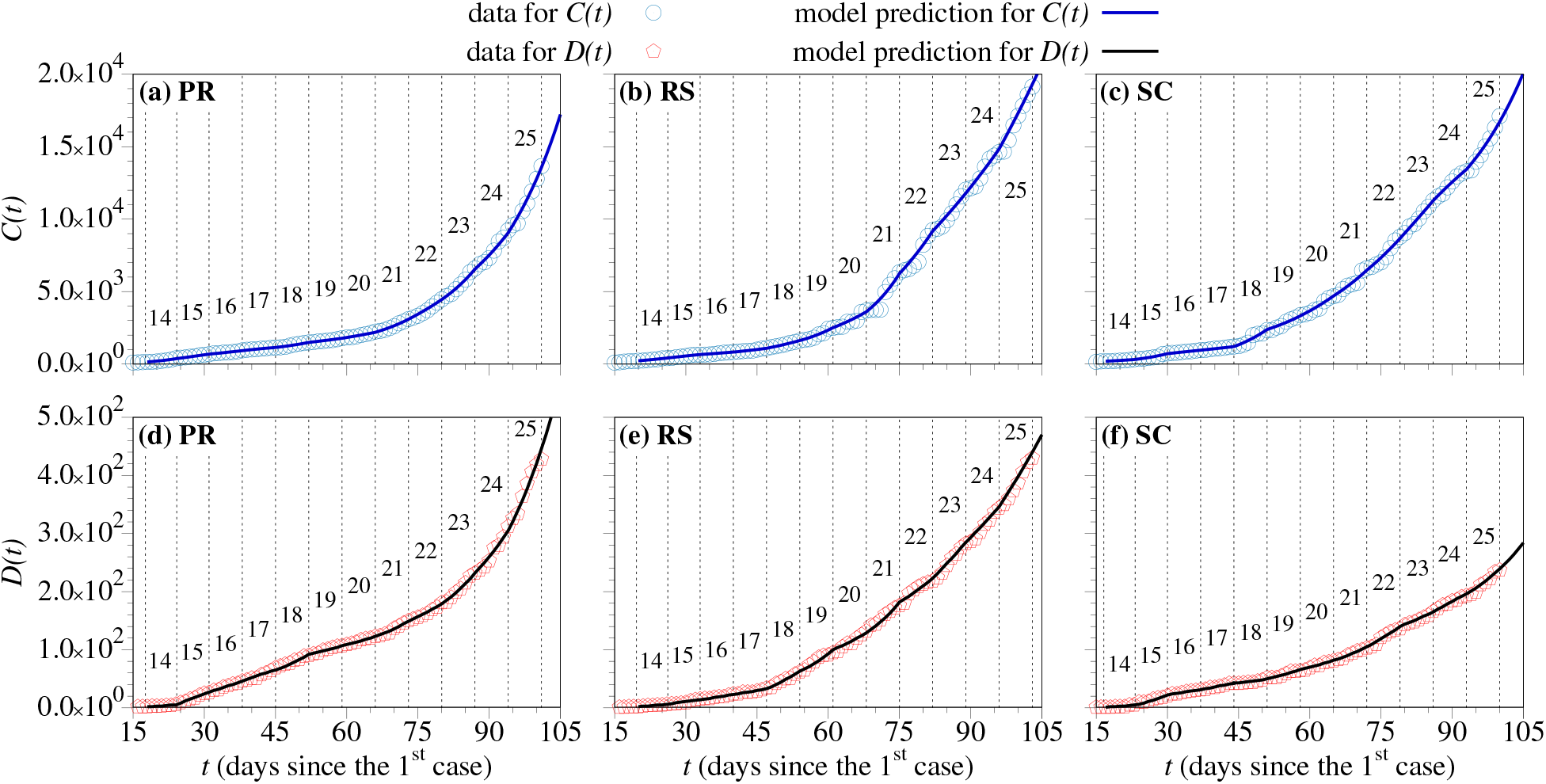
The real data for the cumulative number of confirmed cases (blue circles) and deaths (red pentagons) together with the model predictions for the states of PR [(a) and (d)], RS [(b) and (e)], and SC [(c) and (f)]. The EPI weeks are indicated by the numbers in each panel.

## 3. Numerical results

### 3.1. The Effective Reproduction Number (ERN)

The quantity that determines how fast an infectious disease spreads, considering that a relevant part of the population is no longer susceptible and containment measures were taken, is the *Effective Reproduction Number* (ERN) ℛ_*e*_ [20]. The key aim of the containment measures adopted by the government is to reduce the value of the ERN, with the best scenario taking place when ℛ_*e*_ *<* 1, for which the number of newly infected individuals decrease and the pandemic can be controlled. By using the SIRD model, we can estimate *ℛ*_*e*_(*k*) for each EPI week *k* by the relation [26] 

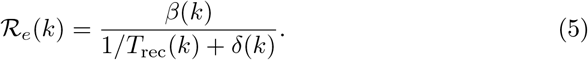

In Fig. 3, we show the values of *ℛ*_*e*_(*k*), for each EPI week *k*, for the states of PR, RS, and SC. We can see that, for the three localities, the ERN oscillates considerably, but remains in most of the time between the values 1 and 3. However, the state of PR keeps *ℛ*_*e*_ ≥ 2 in the last three EPI weeks, which explains why it is a worrisome moment for this state. Despite *ℛ*_*e*_(25) *>* 2 in the last EPI week, SC is the state of the South Region with the lowest value of ⟨*ℛ*_*e*_(*k*)⟩ (see Table 2), which could be a consequence of the implementation of early containment measures. RS, in turn, has the lowest current value for the ERN, *ℛ*_*e*_(25) = 1.25. Although RS is the state of the South Region with the highest number of infected people and deaths (see Table 1), its ERN has been decreasing in the last four weeks.

**Figure 3.**
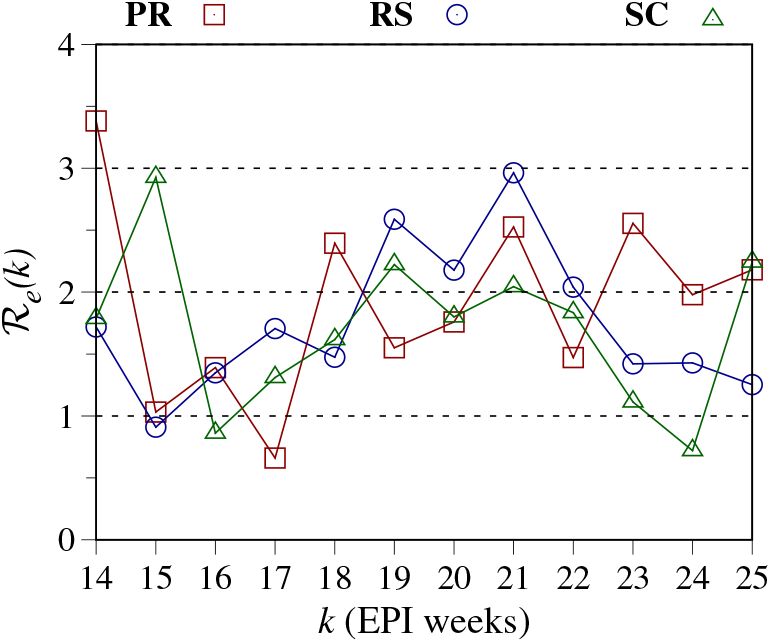
Value of the ERN *ℛ*_*e*_(*k*) for each EPI week *k* for the states of PR, RS, and SC. The mean value of *ℛ*_*e*_(*k*) for each locality is displayed in Table 2.

**Table 2.** Table with the mean values of the epidemiological parameters of the SIRD model. The values of ⟨*β*(*k*)⟩ and ⟨*δ*(*k*)⟩ are given in days^−1^,⟨ *T*_rec_(*k*)⟩ is given in days, and *ℛ*_*e*_ is a dimensionless parameter.

| State | $\langle\beta(k)\rangle$ | $\langle T_{\text{rec}}(k)\rangle$ | $\langle\delta(k)\rangle$ | $\langle\mathcal{R}_e(k)\rangle$ | $\mathcal{R}_e(25)$ |
| --- | --- | --- | --- | --- | --- |
| PR | 0.12 | 17.4 | 0.54% | 1.91 | 2.18 |
| RS | 0.11 | 17.4 | 0.35% | 1.75 | 1.25 |
| SC | 0.11 | 17.0 | 0.22% | 1.71 | 2.24 |

We mention that there are other more accurate techniques to calculate the value of *ℛ*_*e*_ (see, for example, the Refs. [30, 31, 32]). The method used in this work has limitations since it is based on a simple epidemiological model (which does not consider the symptomatic and asymptomatic subgroups of individuals and the incubation period, for example) and depends on a minimum number of experiments to define the parametric set. However, the thorough adjustment procedure and the choice of the parametric intervals, defined according to the real data and studies about the epidemiological and clinical characteristics of COVID-19, allowed us to provide reliable values for the ERN and to capture trends for such quantity. In Table 2, we display the mean values of *β*(*k*), *T*_rec_(*k*), *δ*(*k*), and ℛ_*e*_(*k*) for each state of the Brazilian South Region. These mean values were calculated over all the EPI weeks considered in our analysis. The value of *ℛ*_*e*_(25), obtained for the EPI week number 25, is also displayed since such quantity is used in the next subsection to analyze the trends for the pandemic in each locality.

### 3.2. Flattening the COVID-19 epidemic curve

In Figs. 4(a), 4(b), and 4(c), we show some projections for the cumulative number of confirmed cases *C*(*t*) for the states of PR, RS, and SC, respectively, using the logarithm scale in the vertical axis. When the real data are not available anymore, the blue continuous curves represent the trend for *C*(*t*), obtained by considering the hypothetical scenario for which the value ℛ_*e*_(25) found for each locality remains unchanged until day 150 after the first case. In Figs. 4(a) and 4(c), we can see that for PR and SC, for which ℛ_*e*_(25) *>* 2, the value of *C*(*t*) might keep an exponential increase after the power-law growth showed until then [3, 5]. On the other hand, in Fig. 4(b) it is possible to see that a smoother growth is expected for RS.

**Figure 4.**
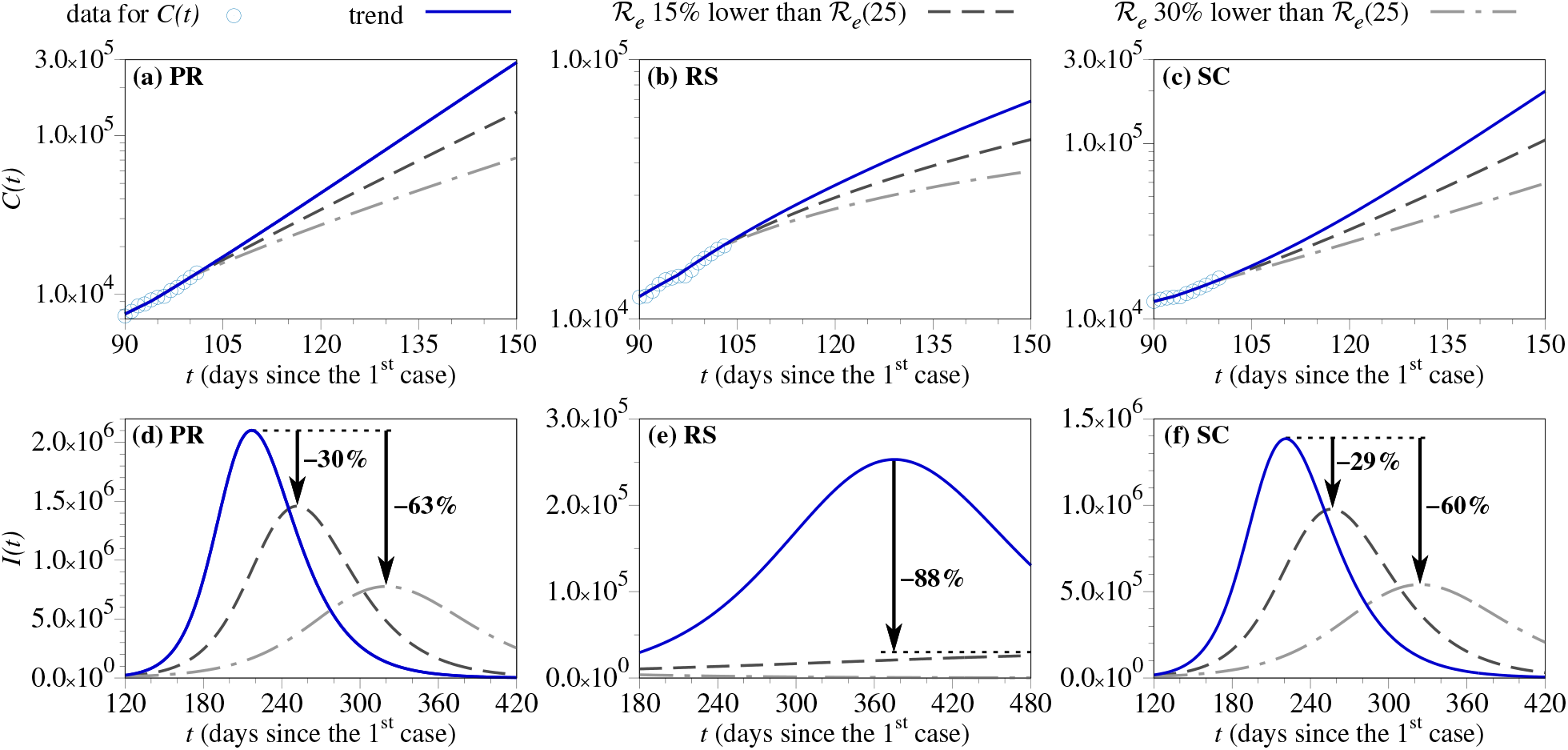
The real data for the cumulative number of confirmed cases *C*(*t*) (blue circles) and its model prediction for the states of PR in (a), RS in (b), and SC in (c). The blue continuous lines are the current trend for each locality, obtained by keeping *ℛ*_*e*_(25) until day 150 after the first case. The dark gray dashed and gray dash-dotted lines represent scenarios obtained by decreasing *ℛ*_*e*_(25) by 15% and 30%, respectively. The same scenarios are shown in the panels (d), (e), and (f) for the number of active infected people *I*(*t*). In these panels, the arrows indicate the percentage reduction in the peak of each scenario when compared with the current trend.

The epidemiological parameters vary daily, and it is not probable that the ERN remains unchanged for a long time. For this reason, long-term projections must be revised whenever a change in the trend occurs. Considering such probably changes, we simulated scenarios in which the ERN decreases in the coming days. As shown in Eq. (5), the ERN is related to the parameters *β, T*_rec_, and *δ*. Since the recovery and death rates depend on several circumstances, including pharmacological treatments, in our simulations we kept fixed these parameters and varied the ERN changing the contact rate *β*(25). This parameter carries all information regarding non-pharmacological measures as, for example, to maintain 6-feet social distancing and to use simple cloth face coverings.

In Fig. 4, the dark gray dashed and gray dash-dotted lines describe projections obtained by decreasing the current contact rate *β*(25) by 15% and 30%, respectively, which means to reduce by 15% and 30% the ERN according to Eq. (5). It is possible to see that a slight decrease in the current ERN might avoid the infection of tens of thousands of people and, consequently, hundreds of deaths. These simulations show the relevance of the containment measures that allow to decrease the contact rate *β* and, as a consequence, to decrease the ERN.

Another important quantity obtained from the SIRD model is the number of active infected people *I*(*t*), plotted in Figs. 4(d), 4(e), and 4(f) for the states of PR, RS, and SC, respectively. The blue continuous curves show the trend for each state, and we can see in Fig. 4(d) that the peak of *I*(*t*) for PR might be the highest of the South Region and also might occur earlier. On the other hand, the curve of *I*(*t*) for RS is the most flattened, which means that the peak of *I*(*t*) expected for this state is lower and might occur later. These results suggest that the flattening of the epidemic curve is related to the current value of ERN, as demonstrated in recent studies [5, 10, 33, 34].

In Figs. 4(d), 4(e), and 4(f), the dark gray dashed curves show that, by decreasing the current ERN by 15%, it is possible to generate a reduction of 30% and 29% in the peak of *I*(*t*) for PR and SC, respectively, while an important shrinkage of 88% occurs for the state of RS. A more efficient flattening of the epidemic curve takes place by decreasing *ℛ*_*e*_(25) by 30% for PR and SC (gray dash-dotted curves). For RS, this reduction results in *ℛ*_*e*_ *<* 1, which means that the peak of *I*(*t*) has already occurred and the gray dash-dotted line is in a descendent way. This result can be explained by analyzing Eqs. (2) and (5). Once *ℛ*_*e*_ *<* 1, then *β <* 1*/T*_rec_ + *δ*, and the number of individuals exiting the subgroup *I* is larger than the number of individuals entering this subgroup.

### 3.3. Gradual mitigation strategies to control the hospital bed occupancy

In the absence of a vaccine, to achieve a reduction in the contact rate between susceptible and infectious individuals by isolating the latter is a critically important strategy that can control the spread of the infection [35]. In this subsection, we propose some mitigation strategies to reduce the contact rate gradually and to avoid the health care system collapse. We made this analysis for the states of PR and RS, but it can be applied to any locality.

According to the epidemiological reports published daily by the State Health Offices, about 8% of the active infected people *I*(*t*) need medical assistance [36, 37]. In our simulations, however, we considered that the hospital bed demand is equal to 10% of *I*(*t*) to obtain a reliable estimation. Furthermore, it is essential to know the current number of hospital beds that each state allocates exclusively to COVID-19 patients. Until June 20^th^, 2020, PR reserved 2027 hospital beds for COVID-19 patients [36]. RS, in its turn, plans to implement 2295 hospital beds in the whole state, according to the contingency plan of the RS’s government [37]. These numbers are indicated in Fig. 5 by the vertical black dashed lines. In this figure, we can see that such quantities of hospital beds are insufficient to supply the demand for both states considering the current trend (blue continuous curves). The only difference between PR and RS is the time for collapse, which might occur earlier in PR. The scenario in which ℛ_*e*_(25) is reduced by 30% is also insufficient for PR, as shown in Fig. 5(a) by the gray dash-dotted line. However, for RS this scenario provides a flattened curve for the hospital bed demand once *ℛ*_*e*_ *<* 1.

**Figure 5.**
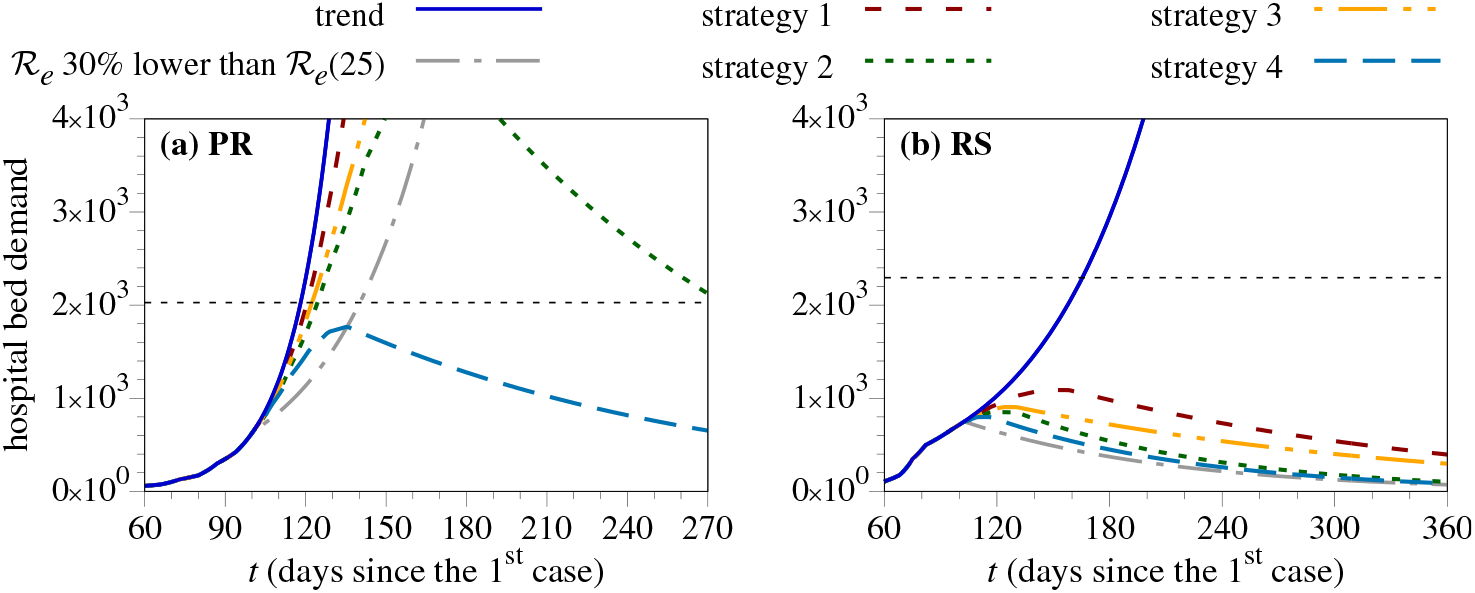
Hospital bed demand for (a) PR and (b) RS, considering that 10% of the active infected people *I*(*t*) will need medical assistance. The vertical black dashed line indicates the number of hospital beds offered for each state.

To avoid overwhelming the health care systems in the coming days, we propose some mitigation strategies that can be applied to an entire community or region, aiming to reduce the personal interactions and thereby the transmission risk. These strategies consist of to adopt different levels of containment measures at specific intervals of time so that we can reduce the contact rate *β* gradually until *ℛ*_*e*_ *<* 1. Such actions can include mild measures as temperature monitoring in public places, the use of cloth face coverings, and the prohibition of group gathering with more than a specific number of people in any public place, until more rigid measures as the cancellation of services and events, the restriction on the entry of foreigners, the control of the entry into and exit from narrowly defined geographic areas, and to restrict people’s transit and business operations, for example [2]. Starting from the current ERN for each state, given in Table 2, the strategies are the following:

i. To reduce by 5% the value of *ℛ*_*e*_ every two weeks until *ℛ*_*e*_ *<* 1. It will take 20 weeks for PR and 8 weeks for RS.
ii. To reduce by 10% the value of *ℛ*_*e*_ every two weeks until *ℛ*_*e*_ *<* 1. It will take 10 weeks for PR and 4 weeks for RS.
iii. To reduce by 5% the value of *ℛ*_*e*_ every week until *ℛ*_*e*_ *<* 1. It will take 10 weeks for PR and 4 weeks for RS.
iv. To reduce by 10% the value of *ℛ*_*e*_ every week until *ℛ*_*e*_ *<* 1. It will take 5 weeks for PR and 2 weeks for RS.

The results obtained by applying these strategies are shown in Fig. 5. For RS, for which *ℛ*_*e*_(25) = 1.25, even the mildest strategy (strategy 1) is efficient to avoid overwhelming the health care system. However, for PR, for which *ℛ*_*e*_(25) = 2.18, only the most rigid strategy (strategy 4) can make the hospital bed demand to be less than the current number offered by the state.

## 4. Conclusions

The SIRD model was used to fit the real data of the cumulative number of confirmed cases and deaths caused by COVID-19 in the states of PR, RS, and SC. Our adjustment procedure consisted of minimizing the RMSE between the model predictions and the real data, as described in Subsection 2.1. By using this procedure, it was possible to reproduce the data from the past, to estimate the ERN for each EPI week, and to make reliable projections for the future.

We found for PR and SC *ℛ*_*e*_(25) *>* 2, and the trend of *C*(*t*) for these localities is to follow an exponential growth, while for the state of RS, for which *ℛ*_*e*_(25) = 1.25, we observed a more attenuated increase for *C*(*t*). We also simulated the effects of reducing the current ERN by applying non-pharmacological measures that reflect on the contact rate *β*. In all cases, to decrease *ℛ*_*e*_ can avoid the infection of tens of thousands of people in the long term.

To avoid overwhelming the health care system, we proposed gradual mitigation strategies that consist of adopting mild or rigid measures at specific intervals of time. For PR, only the most rigid strategy is efficient, while for RS, we verified that mild measures might flatten the epidemic curve when applied gradually. Our results suggest that simple actions as keeping the social distancing and clothing face coverings, when taken early, are efficient to avoid the fast dissemination of the COVID-19.

## Data Availability

The data used in this study are openly available in Ref. [14] of the manuscript

## Acknowledgments

The author thanks Dr. Carlos F.O. Mendes, Dr. Cesar Manchein, MSc. Eduardo L. Brugnago, and Dr. Marcus W. Beims for the fruitful discussions.

## Availability of materials and data

The data used in this study are openly available in Ref. [14].

## References

[1] World Health Organization. Coronavirus disease (COVID-2019) situation reports. https://www.who.int/emergencies/diseases/novel-coronavirus-2019/situation-reports/, 2020.

[2] Centers for Diseases Control and Prevention. Coronavirus Disease (2019). https://www.cdc.gov/coronavirus/2019-nCoV/index.html, 2020.

[3] C. Manchein et al. Strong correlations between power-law growth of COVID-19 in four continents and the inefficiency of soft quarantine strategies. Chaos, 30:041102, 2020.

[4] E. L. Brugnago, R. M. da Silva, C. Manchein, and M. W. Beims. How relevant is the decision of containment measures against COVID-19 applied ahead of time? 2005.01473, 2020.

[5] R. M. Da Silva, C. F. O. Mendes, and C. Manchein. Scrutinizing the heterogeneous spreading of COVID-19 outbreak in Brazilian territory. medRxiv:10.1101/2020.06.05.20123604, 2020.

[6] N. Crokidakis. COVID-19 spreading in Rio de Janeiro, Brazil: Do the policies of social isolation really work? Chaos Soliton. Fract., 136:109930, 2020.

[7] E. Alcantara et al. Investigating spatiotemporal patterns of the COVID-19 in Sao Paulo State, Brazil. medRxiv:10.1101/2020.05.28.20115626, 2020.

[8] L. R. P. de Alcantara et al. Using different epidemiological models to modeling the epidemic dynamics in Brazil. medRxiv:10.1101/2020.04.29.20085100, 2020.

[9] J. T. Wu, K. Leung, and G. M. Leung. Nowcasting and forecasting the potential domestic and international spread of the 2019-nCoV outbreak originating in Wuhan, China: a modelling study. The Lancet, 395:689–697, 2020.

[10] L. Xue et al. A data-driven network model for the emerging COVID-19 epidemics in Wuhan, Toronto and Italy. Math. Biosci., 326:108391, 2020.

[11] D. Fanelli and F. Piazza. Analysis and forecast of COVID-19 spreading in China, Italy and France. Chaos Soliton. Fract., 134:109761, 2020.

[12] E. L. Piccolomini and F. Zama. Preliminary analysis of COVID-19 spread in Italy with an adaptive SEIRD model. 2003.09909, 2020.

[13] F. Ndaïrou, I. Area, J. J. Nieto, and D. F. M. Torres. Mathematical modeling of COVID-19 transmission dynamics with a case study of Wuhan. Chaos Soliton. Fract., 135:109846, 2020.

[14] Ministério da Saúde - Painel Coronavírus. https://covid.saude.gov.br/, 2020.

[15] W. O. Kermack and A. G. McKendrick. A contribution to the mathematical theory of epidemics. Proc. R. Soc. Lond. A, 115(772):700–721, 1927.

[16] D. J. Daley and J. Gani. Epidemic Modelling: An Introduction. Cambridge University Press, 2001.

[17] E. Vynnycky and R. White. An Introduction to Infectious Disease Modelling. Oxford University Press, 2010.

[18] W. O. Kermack and A. G. McKendrick. Contributions to the mathematical theory of epidemics. ii - the problem of endemicity. Proc. R. Soc. Lond. A, 138(834):55–83, 1932.

[19] R. M. Anderson and R. M. May. Infectious Diseases of Humans: Dynamics and Control. Oxford University Press, 1991.

[20] H. W. Hethcote. The mathematics of infectious diseases. SIAM Review, 42(4):599–653, 2000.

[21] S. Ma and Y. Xia. Mathematical understanding of infectious disease dynamics. World Scientific Publishing Company, 2009.

[22] H. Hethcote, W. Wang, and Y. Li. Species coexistence and periodicity in Host-Host-Pathogen models. J. Math. Biol., 51:629–660, 2005.

[23] J. O. Lloyd-Smith, W. M. Getz, and H. V. Westerhoff. Frequency-dependent incidence in models of sexually transmitted diseases: portrayal of pair-based transmission and effects of illness on contact behaviour. Proc. R. Soc. Lond. B, 271(1539):625–634, 2004.

[24] M. Gatto et al. Spread and dynamics of the COVID-19 epidemic in Italy: Effects of emergency containment measures. Proc. Natl. Acad. Sci. U.S.A, 117(19):10484–10491, 2020.

[25] E. D. Sontag. For differential equations with r parameters, 2r + 1 experiments are enough for identification. J. Nonlinear Sci., 12:553, 2002.

[26] C. Anastassopoulou, L. Russo, A. Tsakris, and C. Siettos. Data-based analysis, modelling and forecasting of the COVID-19 outbreak. PLOS ONE, 15(3):1–21, 2020.

[27] S. Chatterjee et al. Studying the progress of COVID-19 outbreak in India using SIRD model. medRxiv:10.1101/2020.05.11.20098681, 2020.

[28] J. H. Beigel et al. Remdesivir for the treatment of Covid-19 — Preliminary Report. N. Engl. J. Med.

[29] R. Lu et al. Epidemiological and clinical characteristics of COVID-19 patients in Nantong, China. J. Infect. Dev. Ctries, 14:440–446, 2020.

[30] T. A. Mellan et al. Report 21: Estimating COVID–19 cases and reproduction number in Brazil. http://hdl.handle.net/10044/1/78872, 2020.

[31] J. Wallinga and P. Teunis. Different Epidemic Curves for Severe Acute Respiratory Syndrome Reveal Similar Impacts of Control Measures. Am. J. Epidemiol., 160(6):509–516, 2004.

[32] R. N. Thompson et al. Improved inference of timevarying reproduction numbers during infectious disease outbreaks. Epidemics, 29:100356, 2019.

[33] Z. Feng, J. W. Glasser, and A. N. Hill. On the benefits of flattening the curve: A perspective. Math. Biosci., 326:108389, 2020.

[34] W. C. Roda, M. B. Varughese, D. Han, and M. Y. Li. Why is it difficult to accurately predict the covid-19 epidemic? Infect. Dis. Model., 5:271–281, 2020.

[35] Z. Ma, Y. Zhou, and J. Wu. Modeling and Dynamics of Infectious Diseases (Series in Contemporary Applied Mathematics). World Scientific Publishing Company, 2009.

[36] Secretaria da Saúde - Estado do PR. Informe Epidemiológico Coronavírus (COVID-19). http://www.saude.pr.gov.br/Pagina/Coronavirus-COVID-19, 2020.

[37] Secretaria da Saúde - Estado do RS. Painel Coronavírus RS. http://ti.saude.rs.gov.br/covid19/, 2020.

